# Evaluating the impact of minimum unit pricing for alcohol on road traffic accidents in Scotland: a controlled interrupted time series study

**DOI:** 10.1101/2022.12.04.22283071

**Authors:** Francesco Manca, Rakshita Parab, Daniel Mackay, Niamh Fitzgerald, Jim Lewsey

## Abstract

**Background:** On 1^st^ May 2018, Scotland implemented Minimum Unit Pricing (MUP) of £0.50 per unit of alcohol to lower alcohol consumption and related harms, and reduce health inequalities. We assessed the impact of MUP on road traffic accidents (RTAs) after 20 months of implementation.

**Methods:** A controlled interrupted time series design was used to evaluate the impact of MUP on RTAs (total, fatal, night-time) in Scotland and any effect modification across socio-economic deprivation groups. RTAs in England and Wales (E&W) were used as a control group. Covariates of severe weather events, bank holidays, seasonal and underlying trends were included.

**Results:** In Scotland, MUP implementation was associated with a 7.2% (95% CI: 0.9%,13.7% P=0.03) increase in the total number of RTAs. For the corresponding period in E&W, there was a 0.9% increase (95% CI: -2.3%,3.2% P=0.75). It is implausible that MUP caused this increase in RTAs, with the most likely explanation of these results being that unmeasured time-varying confounding affected Scotland and E&W differently. There was no evidence of differential impacts of MUP by level of socio-economic deprivation.

**Conclusion:** The introduction of MUP in Scotland was not associated with a lower level of RTAs.

## Introduction

In 2016, alcohol consumption was responsible for more than 5% of all deaths worldwide, causing 5.1% of all disability (DALY) [1]. Drinking harmful levels of alcohol can cause both short- and long-term health risks. While long-term risks are mainly alcohol-related chronic illness and overall societal burdens, short-term risks are related to alcohol misuse and indirectly associated injuries such as violence, homicides, poisonings but also traffic injuries [1]. Further, those experiencing the most socioeconomic disadvantage have the highest levels of alcohol-related harm, which has shown to importantly contribute to inequalities in total mortality in European countries[2]. Alcohol-related harm is high in the UK with alcohol the biggest risk factor for deaths and ill-health among 15-49 years olds and the 5^th^ largest risk factor across all ages [3]. Within the UK, in 2020, Scotland had the highest alcohol-specific death rate of the constituent countries – 21.5 per 100,000 persons, compared to 19.6, 13.9, 13.0 in Northern Ireland, Wales and England, respectively [4].

Following a package of other measures aimed to reduce the high levels of alcohol consumption and subsequent harm in Scotland, the Scottish Government implemented the minimum unit price for alcohol (MUP) on 1^st^ May 2018 [5]. MUP in Scotland is a policy setting a floor price of £0.50 per unit of alcohol (one unit=8g or 10ml of ethanol) below which it cannot be legally sold, thus making alcohol less affordable. As well as reducing overall harms, the aim of MUP is to reduce inequalities by targeting sales of cheap and high-strength alcohol products mainly purchased by the most socioeconomically disadvantaged groups (who have the highest levels of alcohol-related harms [6-8]). Scotland was the first country to implement nationally a homogeneous MUP for alcohol volume in beverages. Therefore, evaluations of MUP in Scotland for a range of outcomes have international relevance as other countries have either subsequently implemented (Republic of Ireland and Wales) or are considering implementation of MUP in the future (England and others).

Evidence on the effectiveness of MUP in Scotland is starting to emerge. It has been shown that, after its first year, MUP was associated with a 3.5% reduction in off-trade alcohol sales per adult [9], with another study using a different data source showing greatest reductions in sales in those households buying the most alcohol [10]. However, MUP was not associated with changes in alcohol-related emergency department visits [11] nor some alcohol-related crimes [12].

It is well established that alcohol use is associated with road traffic accidents (RTAs), with a dose-response relationship between fatal injury and blood alcohol concentration[13]. However, there is only a small evidence base on how minimum alcohol prices are associated with RTAs. A Canadian study[14] observed that increases in provincial minimum alcohol prices were associated with reductions in alcohol-related traffic violations (but not in non-alcohol-related traffic violations). In 2020, a regional report from the Northern Territory of Australia regarding the implementation of MUP at a different extent to Scotland ($1.30 per ‘standard drink’ which is equal to £0.75 per UK unit *-*currency conversion in July 2022) reported a significant instant reduction in the level of alcohol-related RTAs resulting in injury or fatality [15]. In 2021, a study investigating the effect of MUP on RTAs in Scotland [16] found a reduction of 0.28-0.35 fewer daily motor vehicle collisions per million inhabitants (an important reduction considering an average of 3.23 RTAs per million across the study period). However, this study (as well as [15]) have short post-intervention periods (8 months), while it plausible that MUP’s indirect effects on alcohol-related harms have different-size lagged impacts which take longer followup periods to emerge as previously shown for other outcomes and contexts [17, 18]. Further, any differential effects across levels of socioeconomic deprivation (an aim of MUP policy in Scotland) were not considered.

The aim of this paper is to evaluate whether introduction of MUP in Scotland was associated with the level of RTAs in the first 20 months after implementation. Further, we evaluated whether any association varied by level of socioeconomic deprivation and categories of RTAs which have higher likelihoods of being alcohol-related.

## Methods

We used an interrupted time series design to establish whether MUP implementation in Scotland was associated with a variation in the level of RTAs. We assessed the impact of the legislation on the number of total weekly RTAs, before assessing specific subcategories more likely to be alcohol-related events in line with official UK Government figures [19], namely fatal RTAs and night-time (from 6pm to 6am) RTAs. We used the corresponding data for England and Wales (E&W) as a geographical control group. For total RTAs, analyses were repeated for two socio-economic deprivation groups: the most deprived tenth and the rest of the population, as measured by either the Scottish Index of Multiple Deprivation (SIMD) (Scotland) [20] or Index of Multiple Deprivation (IMD) (E&W)[21]. To assess the effect of a policy regarding alcohol pricing on RTAs, data on failed breath tests and drink-driving episodes would comprise an ideal outcome, however, such data has numerous difficulties. For instance, the accuracy of drink-driving data strictly depends on breath tests for non-fatal accidents and from coroner reports for fatal accidents. However, toxicology data coming from coroners are not available for all killed drivers, and they are not accessible for all relevant cases [19], producing high sampling uncertainty around official drink-driving estimates. Further, alcohol consumption by pedestrians is a significant factor in a subset of RTAs which do not involve drink-driving. Therefore, as drink-driving outcomes have these methodological uncertainties, we used data on total RTAs and on specific categories that are more likely to be alcohol-related.

Data on RTAs and casualties for the UK were obtained from the Road safety statistics division at the UK Department for Transport [22]. The dataset contained all personal injury accidents on public roads that were reported to the police [23]. In the dataset, every accident was recorded with the level of severity (from ‘slight’ to ‘fatal’), date and time and with the number of casualties. The data used covers the period 1st Jan 2016 to 31st December 2019, providing 28 months (121 weeks) before the intervention and 20 months (87 weeks) after. The casualties dataset contained a variable on the IMD for RTAs recorded in England or Wales only. For Scotland RTAs, we used the postcode of the casualties to obtain SIMD for each casualty. Whenever RTAs involved more than one person, the lowest socioeconomic deprivation level was used for analysis. Alternative analyses using the highest level of deprivation were also run.

We used weeks as level of data aggregation to remove daily ‘noise’ and multiple seasonalities (weekly and yearly) for easier detection of the trend component of the series. We first ran a descriptive analysis to assess the general trend and patterning of weekly RTAs over time and to detect any outliers. We then used Seasonal Autoregressive Integrated Moving Average models (SARIMA) for inferential analyses. To reduce the impact of outliers, to remove exponential variance and for ease of comparison with other studies, the outcome variable was log-transformed. With log-transformed series, the coefficient of independent variables in the SARIMA models can be approximately interpreted as the percentage variation in the level of RTAs. The effect of MUP was assessed by introducing a binary variable in the SARIMA model, assuming a value of 0 before the week the policy was introduced and 1 after. An underlying pre-intervention deterministic trend variable [24] considering the time elapsed since the start of the study was used as a model covariate. Alternative models with both a change in level and trend were analysed. Different SARIMA models were assessed using a correlogram of the series and after model estimation assessment of white noise of the residuals using portmanteau test [24]. The best fitting model was then selected based on information criteria (Akaike and Bayesian Information Criteria). Models were further adjusted for weeks with severe weather events (collected by the Met Office for the UK [25]) and weeks with bank holidays.

Fatal RTAs had low weekly numbers in both Scotland and E&W. In particular, in Scotland, a few weeks had zero records. Therefore, a commonly used log(x+1) transformation was applied to the series. However, it has been recently shown that results based on this transformation may provide biased estimates [26], therefore alternative sensitivity analyses were used to address this (as described below).

Regarding socioeconomic deprivation group for E&W, there was a relevant difference in the amount of weekly missing data in the period before (326) and after MUP (161) implementation (Table 1). This led to an increase of the overall number of RTAs having level of socioeconomic deprivation recorded from the beginning of the analysis, while, overall, the number of RTAs decreased overtime. These features would limit the meaning of the inferential analyses for the socioeconmic deprivation groups for E&W. For this reason, results on this series are reported mainly for completeness and only for the main analysis. In contrast, missing data on the level of deprivation in Scotland were similarly distributed between pre- and post-intervention periods.

**Table 1.** Average Weekly number of RTAs in Scotland and England & Wales over years and comparison of total and subgroups of RTAs before and after MUP implementation

|  | Scotland |  |  | England and Wales |  |  |
| --- | --- | --- | --- | --- | --- | --- |
|  | Mean (SD) weekly RTAs | Year on Year Difference |  | Mean (SD) weekly RTAs | Year on Year Difference |  |
| 2016 (pre MUP) | 160.50 (18.1) |  |  | 2466.83 (201.2) |  |  |
| 2017 (pre MUP) | 136.79 (16.0) | -23.71 (-15%) |  | 2362.87 (201.5) | -103.96 (-4%) |  |
| 2018 (MUP after 1.5.2018) | 123.17 (17.9) | -13.62 (-10%) |  | 2235.19 (270.0) | -127.68(-5%) |  |
| 2019 (post MUP) | 110.35 (14.4) | -12.82 (-10%) |  | 2163.96 (195.9) | -71.23 (-3%) |  |
|  | Pre MUP<br>1<sup>st</sup> Jan2016-30<sup>th</sup> April2018 | Post MUP<br>1<sup>st</sup> May2018-31<sup>st</sup> Dec2019 | Year on Year<br>Difference | Pre MUP<br>1<sup>st</sup> Jan2016-30<sup>th</sup> April2018 | Post MUP<br>1<sup>st</sup> May2018-31<sup>st</sup> Dec2019 | Year on Year<br>Difference |
| Total RTAs | 143.85 (23.9) | 117.28 (16.7) | -26.5 (-18%) | 2365.26 (247.7) | 2227.2 (223.7) | -138.07 (-6%) |
| Fatal | 2.86 (1.7) | 3.19 (2.1) | 0.33 (+11%) | 28.84 (6.0) | 29.76 (7.1) | 0.91 (3%) |
| Night-time | 39.13 (9.0) | 31.83 (6.9) | -7.30 (-19%) | 400.45 (71.6) | 674.12 (66.3) | -26.33 (-4%) |
| Most deprived tenth group * | 21.33 (4.9) | 16.36 (4.7) | -4.98 (-23%) | 256.88 (36.8) | 262.25 (28.4) | 5.37 (+2%) |
| 2<sup>nd</sup>-10<sup>th</sup> deprived group * | 110.91 (19.4) | 83.68 (16.3) | -27.23 (-25%) | 1694.53 (221.6) | 1717.72 (189.9) | 23.19 (+1%) |

\* = Missing data for Scotland were 1634 (13 per week on average) pre intervention and 1006 (12 per week on average) post intervention for Scotland. In England and Wales missing data were 39388 (326 per week on average) pre intervention and 14064 post intervention (161 per week on average).

To compare our findings with previous evidence on the effect of MUP on RTAs [16], we reproduced our analysis with similar data using a shorter post-intervention period ending in December 2018. However, we still started our time series in 2016 and used different time units (weekly).

### Sensitivity analysis

For statistically significant results, falsification tests simulating an intervention one year before and one year after the actual date of MUP implementation were evaluated. Two alternative analyses for RTAs concerning fatalities were considered to account for an excess of zeros and a general low number of events. Specifically, a different transformation with Inverse hyperbolic sine transformation (IHS) with small-sample bias correction [26] and a GLM negative binomial model, specifically used to deal with counting data, were also employed and compared with the main analysis.

Finally, we considered models of the difference between the series, *log(RTAs in E&W)-log(RTAs in Scotland)*, as the outcome. If the control series (E&W) have a common trend with the series receiving the policy before the policy introduction, then this model would produce a difference-in-differences type causal estimate directly accounting for the control [27].

## Results

The average weekly number of RTAs for each year and the total number of RTAs before and after MUP implementation are shown in Table 1 in both intervention and control regions. The weekly number of RTAs in Scotland and E&W between 1st January 2016 and 31st December 2019 is shown in Figure1. There is a seasonal trend showing a decrease in the average weekly number of total RTAs in March and April, where the average weekly RTA count was 122.8 and 2111.6 in Scotland and E&W throughout the analysis period, while in the rest of the year was 134.6 (+9.6%) and 2349.6 (+11.3%). Similar seasonal patterns of different strenght are also observed in the series related to night-time RTAs. In contrast, the volume of fatal RTAs in Scotland was too small to detect any seasonal pattern (Figure 1, panel c). The level of weekly RTA was consistently higher in the period before MUP in both intervention and control groups for total RTAs and most of the subcategories (Table 1), except for fatal RTA. Nevertheless, it is easy to identify a declining trend within both pre and post MUP periods for both intervention and control groups. Again, while this tendency is common to most of the subcategories, for fatal RTAs there was an increasing trend over time in Scotland: +0.7 (+20%) pre MUP and +0.5 (+15%) post MUP. For E&W the pattern of the differences was less distinct within the two periods.

**Figure 1.**
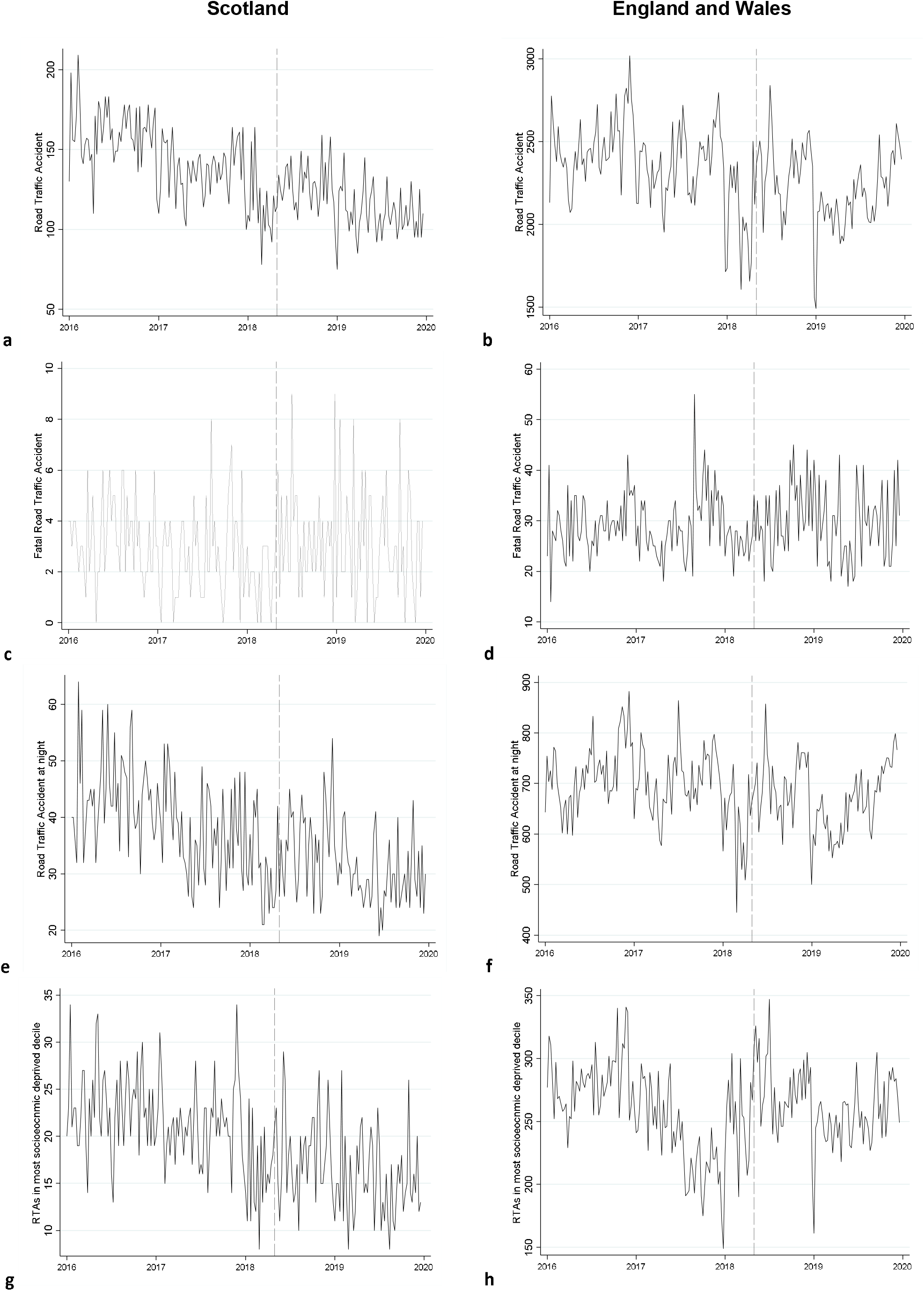
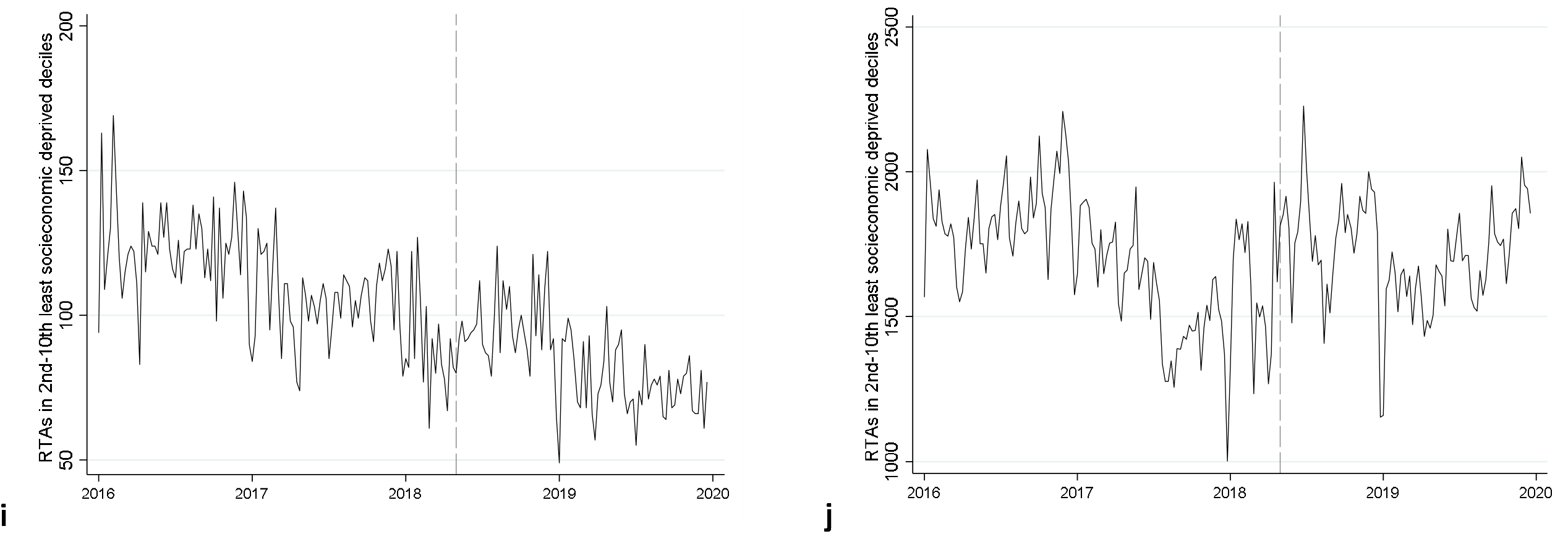
Weekly total RTAs for Scotland (a) and England and Wales (b), weekly fatal RTAs in Scotland (c) and England and Wales (d), weekly night-time RTAs in Scotland (e) and England and Wales (f), weekly total RTAs in most socio-economically deprived group (highest tenth of SIMD/IMD) in Scotland (g) and England and Wales (h), weekly total RTAs in all other socio-economically deprived groups in Scotland (i) and England and Wales (j) between 1st January 2016 and 31st December 2019. Dash vertical line represents date of MUP implementation.

Figure 2 visually describes the percentage changes resulting from the inferential analysis, with full model outputs presented in the supplementary materials. Based on information criteria, models including a change in level only were selected for base case analysis. In Scotland, the introduction of MUP was associated with a 7.2% (95%CI: 0.9%, 13.7% P=0.03) increase in the total number of RTAs. For the corresponding period, in E&W there was a 0.9% increase (95% CI: -2.3%, 3.2% P=0.75). Fatal RTAs had a significant increase in Scotland after MUP of 40.5%, but both Scotland and E&W results for this outcome had considerable uncertainty (see wide confidence intervals). RTAs at night were not significantly associated with MUP introduction in either Scotland or E&W. There was no significant association between the level of RTAs and the introduction of MUP in Scotland for the most socioeconomically deprived tenth and for the 2^nd^-10^th^ deprived tenth groups. The underlying trend was negative in all the models indicating a decreasing pattern of all series over time, and it was always statistically significant, except for the model regarding fatal RTA in E&W (see supplementary material).

**Figure 2.**
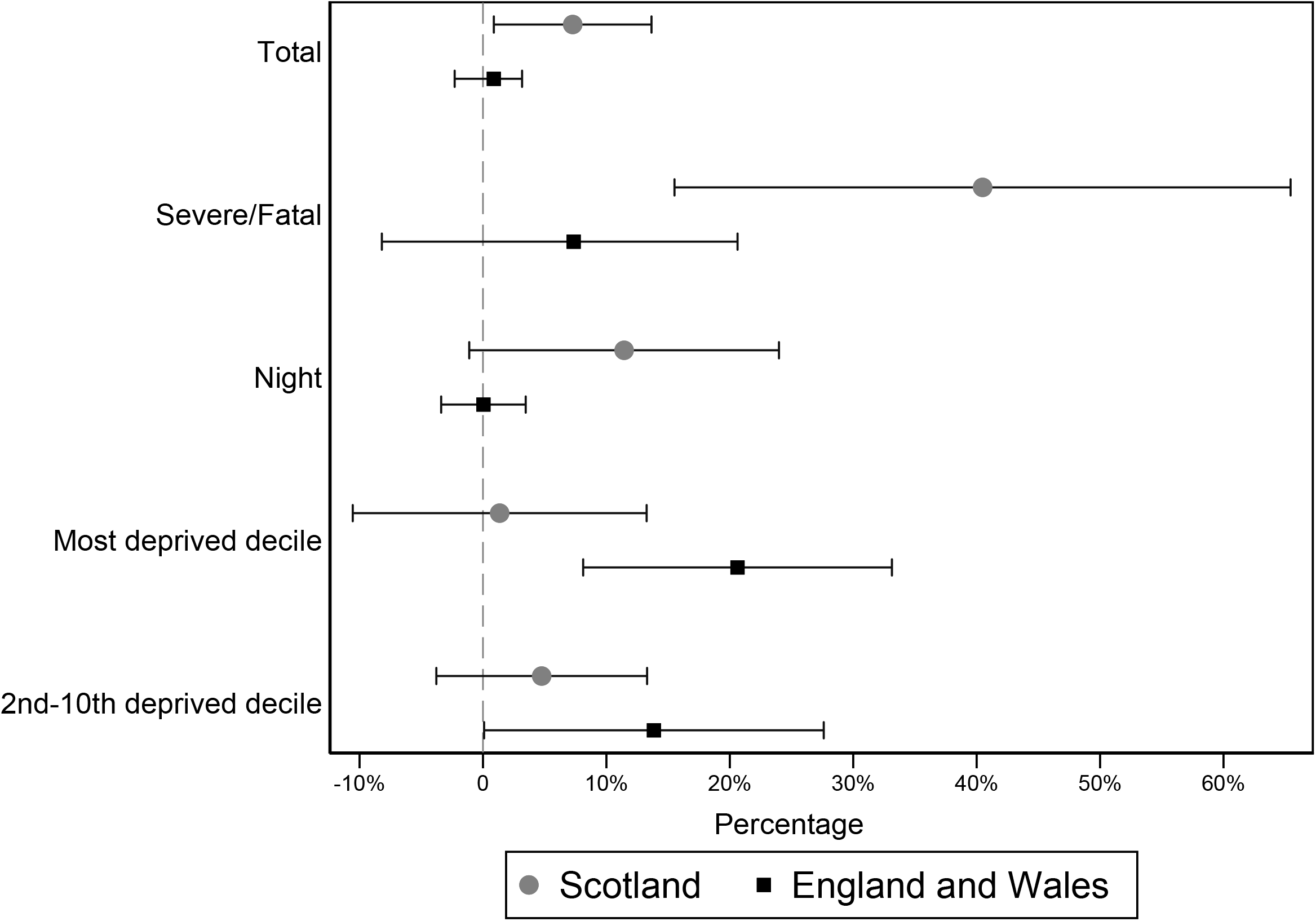

By reproducing a similar analysis to [16] with 8 months post-intervention follow-up, Scotland was associated with a significant relative increase in total RTAs of almost 10%, with a corresponding increase of 5.8% in E&W.

### Sensitivity analysis

Falsification tests anticipating or delaying the implementation of MUP by one year produced results that were not statistically significant for both total and fatal RTAs in both intervention groups, confirming results in the main analysis.

Concerning fatal RTAs, IHS transformation with small sample correction produced models with associated increases of 45% and 4% for the period after MUP implementation for Scotland and E&W, respectively. In line with these results, the negative binomial model for Scotland provided a significant increase of 53% in the incident rate ratio after MUP introduction. While all these alternative models vary the point estimates related to MUP introduction, they did not change the direction of the association.

As the difference in the log of fatal RTAs between the two intervention groups was the only series satisfying common trend requirements (see supplementary materials), we performed the analysis on the difference for this group only. This analysis detected a positive increase in fatal RTAs after MUP introduction across all the three different transformations (log(x+1), IHInverse hS and Poisson regression) - see supplementary materials – confirming results in the main analysis.

## Discussion

This study does not provide evidence that the introduction of MUP was associated with a reduction of RTAs in Scotland for the first 20 months of its implementation, nor that any MUP effect varied by level of socioeconomic deprivation. In fact, the introduction of MUP in Scotland was associated with an increase in total and fatal RTAs that was not observed in the control group of E&W. In our subgroup analysis, we did not find differential MUP effects on RTAs across socioeconomic deprivation groups in Scotland. Regarding fatal RTAs, the low number of weekly observations together with a likely floor effect may have produced high uncertainty in the estimates and consequent low usefulness of results to draw conclusions. However, different transformations and models taking into account potential floor effects detected a significant positive association between MUP and fatal RTAs in line with the main analysis, yet still with high uncertainty (39%, 95% CI: 3.7,74.9 P=0.03). We believe that our findings underlie associations rather than causality. Indeed, it seems implausible that introduction of MUP is causally linked to increased levels of total RTAs, as alcohol sales fell following the introduction of MUP, and alcohol is a significant cause of RTAs. The most likely explanation of these results is that unmeasured time-varying confounding is at differential levels between Scotland and E&W. Evidence for this is seen in the non-parallel trends observed in the pre-MUP parts of the time series shown in Figure 1. Such time-varying confounding could be due to weather, road quality and demographic changes.

We designed a natural experiment, using Scotland as the intervention group and E&W as a concurrent control group. A priori, we believed we chose the best available counterfactual according to theoretical considerations, with the intervention and control groups both part of the UK and likely to have similar underlying temporal trends in RTAs. Also, a large nationally representative dataset was used allowing for a long follow-up and consequently high statistical precision. A potential limitation is that we did not measure drink-drive RTAs directly, and that not all RTAs are reported to the police. However, it is worth noting that both this and other possible time-varying confounders would need to be different between Scotland and E&W to be a source of bias in our analyses.

Overall, according to the economic theory, the increase in the price of alcohol due to MUP could have led to expectations of a decrease in RTAs. These prospects could come from a knock-on effect originated by lowering consumption and in particular the consumption of more harmful and risky drinkers (mainly targeted by the policy [6, 8]). This would have reduced drink-driving behaviours (and increased pedestrian road safety awareness) and, consequently lowered RTAs. However, even by selecting subcategories of RTAs more likely to be affected by alcohol consumption, we did not find any association with a decrease in the number of RTAs. One possible explanation is that a minimum price of £0.50 could be too low to generate such effect with visible repercussions on drink-driving/pedestrian road safety and then in the total number of RTAs. In support of this hypothesis, only 13% of RTAs with fatalities were linked to drink-driving episodes in 2019 [19], and small variations to this may be hard to detectin overall RTAs. Moreover, the general reduction in alcohol consumption due to MUP [9], may not have sufficiently diminuished consumption in those more lilkely to be drink drive offenders. For instance people with alcohol dependence, if they drink-drive, tend to do so at well above the drink-drive limit, and are not the main group who may benefit from MUP[28]. Further, MUP did not affect all alcohol on sale: prices in pubs and restaurants that typically were already above the floor price of £0.50 did not increase. Another explanation could be that even if RTAs affected by alcohol drinking were theoretically affected by MUP, the period to assess such changes in our analyses (20 months) was too short to change certain drinking attitudes and population behaviours

Additionally, national statistics for Great Britain [19] show a sharper decrease in drink-driving accidents than in all other RTAs, suggesting that some environmental or behavioral factors in the population may act as confounders in both intervention and control areas by generating this RTA reduction over time. In this scenario, our analysis already picking an overall decreasing trend may then associate a variation of this pattern, such as a deceleration of a decrease in total RTAs, or a different change between the two series (alcohol-related RTAs vs all other RTAs) with MUP.

Our results are in contrast with those already published [15, 16, 18]. In the previous evaluation of MUP in Scotland on RTAs [16], the authors found that RTAs increased in 2018 and by showing that Scotland had a lower growth than E&W, associated this relative decrease to MUP. Using a longer time frame (adding 2 years before intervention and one after), weekly data, a different study design and accounting for underlying trends, we found an increase (rather than decrease) from our inferential models in RTAs after MUP. In our analysis, the overall trend over the years was negative (rather than positive) and evaluating the pre-intervention period from 2016, the two groups did not present parallel trends in weekly data (see supplementary material) for overall RTAs. When we reduced the post intervention period to emulate the previous evaluation, our results did not change, which could be due to a longer pre-intervention period being modelled. This could suggest that the previous study may have captured only a momentary trend over time and associated this temporary change between the two intervention groups (maybe related to different seasonality) to MUP. Indeed, they concluded that the effect of MUP implementation may have generated 1.52–1.90 fewer daily collisions in Scotland which, based on our weekly figures (Table 1), is a decrease of 7.4-9.2% (a substantial effect considering that overall MUP was associated with a 3.5% reduction in alcohol consumption [9]). Additionally, the short follow-up period of the previous evaluation may suggest an instant effect of MUP, while similar studies indicate lagged consequences [18]. We believe that by analysing a longer pre- and post-intervention period as well as considering seasonality and autoregressive components, we have provided a more robust analysis. Sherk et al [18], focusing on emergency department (ED) visits rather than on RTAs, showed that raising minimum alcohol pricing in Saskatchewan, Canada was associated to a lagged decrease in motor vehicle-collision-related ED visits only for a certain subcategory of patients (women aged over 25 years). No significant association was found for many other categories of RTA related ED visits subdivided by sex and age. The authors reported that their main hypothesis of a decrease of ED visits due to a raise in minimum alcohol pricing was not substantiated by their findings.

## Conclusion

After 20 months of implementation, there is no evidence of a decrease in RTAs as a consequence of MUP implementation in Scotland. Further, there is no evidence of differential effects by level of socioeconomic deprivation.

## Data Availability

Manuscript data was obtained from the Road safety statistics division at the UK Department for Transport

