## supplementary material for "Evaluating the impact of minimum unit pricing for alcohol on road traffic accidents in Scotland: a controlled interrupted time series study"

### **Regression output for time trend and MUP level.**

#### **ALL RTAs**

```
Scotland
time |  -.0028724   .0002691   -10.67   0.000    -.0033999   -.0023449
 MUP |   .0727278   .0325667     2.23   0.026     .0088982    .1365574
E&W
time |    -.00101   .0001148    -8.79   0.000    -.0012351   -.0007849
 MUP |   .0088023   .0142841     0.62   0.538    -.0191941    .0367986
```

#### **FATAL RTAs**

```
Scotland
time |  -.0035044    .001017    -3.45   0.001    -.0054976   -.0015111
 MUP |   .4048462   .1273429     3.18   0.001     .1552586    .6544338
E&W
  time |  -.0003246   .0005913    -0.55   0.583    -.0014835    .0008343
   MUP |   .0621703   .0735256     0.85   0.398    -.0819372    .2062778
```

#### **NIGHT**

```
Scotland
time |  -.0030994    .000526    -5.89   0.000    -.0041303   -.0020686
 MUP |   .1143684   .0640193     1.79   0.074    -.0111071     .239844
E&W
time|  -.0006315   .0001422    -4.44   0.000    -.0009103   -.0003528
MUP |   .0003779   .0174683     0.02   0.983    -.0338595    .0346152
```

#### **MOST DEPRIVED tenth**

```
Scotland
MUP |   .0135407   .0607525     0.22   0.824     -.105532    .1326135
time |  -.0028894   .0005436    -5.32   0.000    -.0039548    -.001824
E&W
MUP |   .1411663   .0695341     2.03   0.042     .0048819    .2774506
time |  -.0012837   .0006545    -1.96   0.050    -.0025665   -1.00e-06
```

#### **2nd-10th deprived groups**

```
Scotland
  MUP |   .0387297   .0601993     0.64   0.520    -.0792587     .156718
 time |  -.0032108   .0005294    -6.07   0.000    -.0042483   -.0021732
E&W
MUP |   .1411663   .0695341     2.03   0.042     .0048819    .2774506
time |  -.0012837   .0006545    -1.96   0.050    -.0025665   -1.00e-06
```

### **FATAL RTAs TRANSFORMATIONS**

## **l(x+1)**

```
Scotland
time |  -.0035044    .001017    -3.45   0.001    -.0054976   -.0015111
 MUP |   .4048462   .1273429     3.18   0.001     .1552586    .6544338
E&W
time |    -.00101   .0001148    -8.79   0.000    -.0012351   -.0007849
 MUP |   .0088023   .0142841     0.62   0.538    -.0191941    .0367986
```

#### **Inverse hyperbolic sine transformation**

```
Scotland
time |  -.0043927   .0008764    -5.01   0.000    -.0061105    -.00267
 MUP |   .4283237   .1091315     3.92   0.000       .21443    .6422174
E&W
time |  -.0004284   .0006272    -0.68   0.495    -.0016577    .0008009
 MUP |   .0772106   .0766473     1.01   0.314    -.0730153    .2274365
```

#### **Negative binomial (results in incident rate ratio)**

```
Scotland
time |   .9961094   .0013733    -2.83   0.005     .9934214    .9988046
 MUP |   1.531423   .2629018     2.48   0.013     1.093876    2.143987
E&W
time |   .9994033   .0004567    -1.31   0.192     .9985085    1.000299
 MUP |   1.095184   .0583919     1.71   0.088     .9865151    1.215824
```

### **Falsification tests**

**-1yr**

```
Scotland accidents
time |  -.0021575   .0002365    -9.12   0.000    -.0026212   -.0016939
MUP |  -.0279746   .0317351    -0.88   0.378    -.0901743     .034225
E&W
time |  -.0007869   .0001037    -7.59   0.000    -.0009901   -.0005836
 MUP |  -.0228329   .0128113    -1.78   0.075    -.0479426    .0022768
Scotland fatal
time |  -.0001039   .0009281    -0.11   0.911    -.0019229    .0017152
 MUP |  -.0833133    .124863    -0.67   0.505    -.3280402
England fatal
time |  -.0002229   .0005293    -0.42   0.674    -.0012602    .0008145
 MUP |   .0533282   .0729913     0.73   0.465    -.0897321
Scotland night
time |  -.0016437   .0004601    -3.57   0.000    -.0025455    -.000742
 MUP |   -.097068   .0612869    -1.58   0.113    -.2171881
England night
time |  -.0004439   .0001457    -3.05   0.002    -.0007295   -.0001582
 MUP |  -.0282635    .016983    -1.66   0.096    -.0615495    .0050226
Scotland most socioeconomically deprived decile
 MUP |  -.0072268   .0661743    -0.11   0.913     -.136926    .1224724
time |  -.0027531    .000538    -5.12   0.000    -.0038075   -.0016987
Scotland least deprived deciles
 MUP |   .0566446   .0397588     1.42   0.154    -.0212812    .1345703
time |  -.0032398   .0002676   -12.11   0.000    -.0037642   -.0027154
```

**+1yr**
**Scotland accidents**
time | -.0023773	.0001873	-12.70	0.000	-.0027443	-.0020103

```
 MUP |   .0097396   .0314592     0.31   0.757    -.0519192    .0713984
England
time |  -.0010187   .0000662   -15.38   0.000    -.0011486   -.0008889
 MUP |   .0241797   .0155618     1.55   0.120     -.006321    .0546804
Scotland fatal
time |  -.0007773   .0007249    -1.07    0.284   -.00219   .0006435
 MUP |   .0330323   .1079674    0.31     0.760   -.17858   .2446445
England fatal
time |   .0003244   .0003436    0.94   0.345     -.0003491   .0009979
 MUP |  -.0537606   .0588983   -0.91   0.361     -.1691991   .0616779
Scotland at night
time |  -.0022395   .0003867   -5.79   0.000      .0029973   -.0014817
 MUP |  -.0091081   .0638626   -0.14   0.887      -.1342764   .1160602
England night
time |  -.0007266     .00012   -6.05   0.000      -.0009619    .0004914
 MUP |   .0611433   .0247027    2.48   0.013       .0127268    .1095598
Scotland most deprived
 MUP |  -.0350607   .0580873    -0.60   0.546    -.1489097    .0787883
time |  -.0026511   .0004287    -6.18   0.000    -.0034914   -.0018108
Scotland 2nd-10th deprived groups
MUP  |  -.0610501   .0429061    -1.42   0.155    -.1451445    .0230443
time |  -.0026543   .0002161   -12.28   0.000    -.0030778   -.0022308
```

### **Trend difference (pre intervention period)**

#### **total accidents**

```
ldiff_acc    |
time |   .0019429   .0003575     5.43  0.000     .0012422    .0026435
cons|   2.691774   .0260066   103.50   0.000     2.640802    2.742746
```

**Total at night**
ldiff_accnight |

```
time |   .0025508    .000596     4.28   0.000     .0013827    .0037188
cons |   2.748348   .0436364    62.98   0.000     2.662823    2.833874
```

#### **Total severe**

```
ldiff_accsev |
time |   .0041735    .001333     3.13   0.020     .0015608    .0067862
cons |   1.883069   .1020019    18.46   0.000     1.683149    2.082989
```

### **Regressions on difference (fatal RTAs)**

```
Difference
-------------------------------------------------------------------------------
              |             Semirobust
  diff_accsev | Coefficient  std. err.      z    P>|z|     [95% conf. interval]
--------------+----------------------------------------------------------------
diff_accsev   |
ext_weather_E |   .2605986   .1832546     1.42   0.155    -.0985739     .619771
  ext_weather |  -.7768283   .1635984    -4.75   0.000    -1.097475   -.4561814
     ext_snow |   1.337967    .398614     3.36   0.001     .5566976    2.119236
          MUP |  -.3770282   .1507309    -2.50   0.012    -.6724553   -.0816012
         time |   .0035315   .0011948     2.96   0.003     .0011898    .0058732
        _cons |   1.918459   .0817052    23.48   0.000     1.758319    2.078598
Inverse hyperbolic sine transformation
-------------------------------------------------------------------------------
              |             Semirobust
   diffIHSsev | Coefficient  std. err.      z    P>|z|     [95% conf. interval]
--------------+----------------------------------------------------------------
diffIHSsev    |
ext_weather_E |  -.1283322   .2200481    -0.58   0.560    -.5596185    .3029541
  ext_weather |   1.035207   .1971685     5.25   0.000     .6487642     1.42165
     ext_snow |  -1.564516   .3177225    -4.92   0.000    -2.187241   -.9417914
          MUP |   .4311978   .1895179     2.28   0.023     .0597496    .8026459
         time |   -.004209   .0015449    -2.72   0.006    -.0072369   -.0011811
        _cons |  -2.158514   .1116823   -19.33   0.000    -2.377407   -1.939621
--------------+----------------------------------------------------------------
Negative binomial
-------------------------------------------------------------------------------
              |               Robust
         diff |        IRR   std. err.      z    P>|z|     [95% conf. interval]
--------------+----------------------------------------------------------------
         time |   1.000368   .0005545     0.66   0.507     .9992814    1.001455
ext_weather_E |   1.220287   .1310794     1.85   0.064     .9886182    1.506244
  ext_weather |   .5235373   .0566897    -5.98   0.000     .4234263    .6473176
     ext_snow |   .8703801    .032088    -3.77   0.000     .8097072    .9355994
          MUP |   1.001966   .0679575     0.03   0.977     .8772453    1.144419
        _cons |   25.35933   .9926098    82.60   0.000      23.4866    27.38138
-------------------------------------------------------------------------------
```
